# Machine Learning to Predict-Then-Optimize Elective Orthopaedic Surgery Scheduling Improves Operating Room Utilization

**DOI:** 10.1101/2024.08.10.24311370

**Authors:** Johnathan R. Lex, Jacob Mosseri, Jay Toor, Aazad Abbas, Michael Simone, Bheeshma Ravi, Cari Whyne, Elias B. Khalil

## Abstract

**Objective:** To determine the potential for improving elective surgery scheduling for total knee and hip arthroplasty (TKA and THA, respectively) by utilizing a two-stage approach that incorporates machine learning (ML) prediction of the duration of surgery (DOS) with scheduling optimization.

**Materials and Methods:** Two ML models (for TKA and THA) were trained to predict DOS using patient factors based on 302,490 and 196,942 examples, respectively, from a large international database. Three optimization formulations based on varying surgeon flexibility were compared: Any: surgeons could operate in any operating room at any time, Split: limitation of two surgeons per operating room per day and MSSP: limit of one surgeon per operating room per day. Two years of daily scheduling simulations were performed for each optimization problem using ML-prediction or mean DOS over a range of schedule parameters. Constraints and resources were based on a high-volume arthroplasty hospital in Canada.

**Results:** The Any scheduling formulation performed significantly worse than the Split and MSSP formulations with respect to overtime and underutilization (p<0.001). The latter two problems performed similarly (p>0.05) over most schedule parameters. The ML-prediction schedules outperformed those generated using a mean DOS over all schedule parameters, with overtime reduced on average by 300 to 500 minutes per week. Using a 15-minute schedule granularity with a wait list pool of minimum 1 month generated the best schedules.

**Conclusion:** Assuming a full waiting list, optimizing an individual surgeon’s elective operating room time using an ML-assisted predict-then-optimize scheduling system improves overall operating room efficiency, significantly decreasing overtime.

**Highlights:**

- A new approach for elective surgery scheduling was developed combining machine learning prediction of duration of surgery with integer linear programming optimization.
- Models developed on six years of prospective multi-institution data and prediction results were used to generate two years of weekly simulated operating room schedules.
- Three optimization models with varying levels of surgeon constraint were compared, and the model that optimized an individual surgeon’s wait list performed the best.
- Using this approach reduced overtime by 300-500 minutes per week across five operating rooms.

## 1 Introduction

Total knee and hip arthroplasty (TKA and THA, respectively) are the gold-standard treatment for end-stage arthritis of the hip and knee joints. These procedures are the first and second most frequently performed in the US, excluding maternal and neonatal procedures ^1^. In 2018, 1.3 million of these procedures were performed in the US, a 21.9% increase from 2008^1^. This number will continue to increase globally due to an aging population and ongoing obesity pandemic ^2,3^. The ubiquity of these procedures correlates strongly with their burden on healthcare systems globally. In the US, approximately 5% of the gross domestic product (GDP) is to care for musculoskeletal conditions ^4,5^. Despite this spending, wait times for elective surgical procedures in OECD countries continue to increase, conferring extended periods of time with poor quality of life for TKA and THA patients ^6,7^. For these reasons, there is a growing interest and research into improving the efficiency and cost-effectiveness of arthritis care ^8,9^.

Most research has focused purely on the prediction of duration of surgery (DOS) or the optimization of operating room scheduling, ignoring their inherent interrelation ^10–14^. More recently, these approaches have been combined to plan post-surgical beds and plan emergency surgeries based on predicted priority with varying results ^15,16^. DOS prediction models typically represent a large volume of varying procedures, spanning multiple surgical specialties with limited practical applications. Machine learning (ML) models have been applied to predict DOS for various surgeries ^10,12,17–21^. However, in practice, mean time or surgeon-specific rolling mean time is typically used to generate schedules at the operational level.

Research evaluating the optimization of surgery scheduling has been performed using an average or a randomly sampled (typically from a lognormal distribution) DOS value prior to optimizing a schedule through integer linear programming based on either the multiple knapsack or job-shop scheduling problem ^22–24^. Stochastic programming and distributionally robust optimization have also been attempted in order to mitigate the effects of an uncertain DOS, but these approaches assume a distribution of DOS rather than using specific features to aid in prediction ^25^.

To our knowledge, no prior work has combined patient-level DOS predictions with schedule optimization to create an optimized surgical schedule at the operational level. It is known that neural network’s are strong predictors of DOS, however, their realizable improvements when implemented over various surgical schedule optimization problems, while performing simulations accounting for real-world constraints, remains unclear ^21^. The primary aim of this paper was to determine if a two-stage approach utilizing a ML model for prediction of DOS paired with schedule optimization (considering three distinct scheduling formulations) improves operating room over/underutilization compared to using the mean DOS. Secondary objectives were to determine the effect of schedule granularity and length of surgeon wait list on scheduling accuracy.

## 2 Materials and Methods

### 2.1 Setting

Institutional research ethics board approval was obtained for this study (Sunnybrook Health Sciences Centre, Project REB ID #4899). Population level data from the American College of Surgeons National Surgical Quality Improvement (ACS NSQIP) database was utilized to generate our prediction models. This database compiles patient and outcome data following surgery from over 700 hospitals in North America, capturing over 1 million surgeries per year, with a high level of accuracy^26^. The database was queried for all TKA and THA surgical procedures performed between 2014 and 2019. The actual DOS times as reported in the ACS NSQIP dataset were used to inform simulated daily OR schedules. A flowchart that helps visualize the overall two-stage approach is shown in Figure 1.

**Figure 1.**
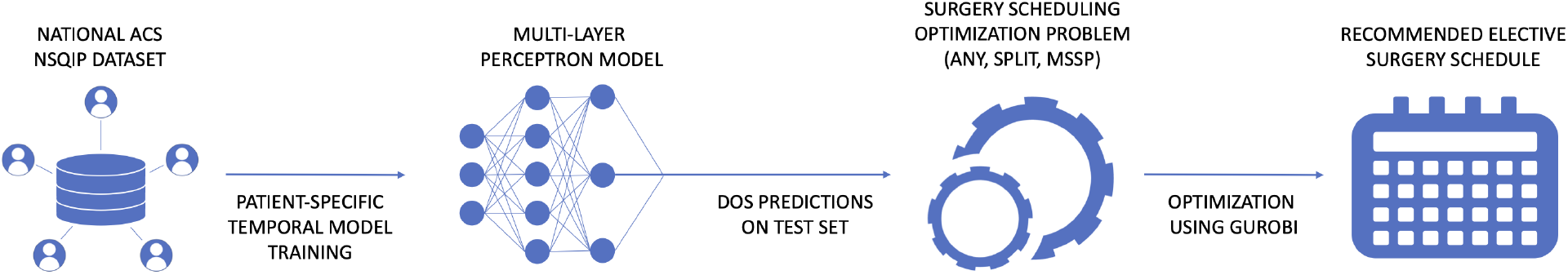
High-level overview of the predict-then-optimize approach. ACS NSQIP = American College of Surgeons National Surgical Quality Improvement Program; DOS = Duration of surgery; MSSP = Multiple Subset Sum Problem

### 2.2 Duration of Surgery Prediction

Data pertaining to 302,490 TKA and 196,942 THA procedures performed during this period as identified from the ACS NSQIP database were used to train models to predict DOS. Abbas et al. identified a PyTorch multilayer perceptron (MLP) model outperformed 10 alternative ML models for predicting DOS for TKA and THA ^21^. Both models were trained on procedures from 2014 to 2017, hyperparameter tuned with Ray on procedures from 2018 and evaluated on procedures from 2019^21,27,28^. These models were generated using the Niagara supercomputer at the SciNet HPC Consortium ^29^. Prediction of DOS from the test subsets of data were used in the optimization model.

### 2.3 Schedule Optimization Formulations

#### 2.3.1 Assumptions and simulation details

The generated scheduling model considered elective case scheduling, in which surgeries are planned for in advance. The constraints and available resources for the model were generated using real-world constraints from the authors’ Institution, the highest-volume elective arthroplasty hospital in Canada. This included cleaning time (30 minutes), number of operating rooms (*n* = 5), number of surgeons (*n* = 11) and days a surgeon was unavailable in a week. The planning horizon of one week (5 workdays) was also based on these constraints. The penalty for an individual OR running overtime (after 5pm), *λ*, was chosen to be double the value of daytime OR underutilization. This was due to the approximate additional costs associated with OR staff working after-hours. The following assumptions regarding scheduling were made: surgeons are available to operate any day anytime except for 0-2 randomly selected days per week per surgeon, and once a surgery is assigned to a surgeon, they must perform it (i.e., no sharing patients). These assumptions were made as the study did not have access to historical data on surgeon availability from the authors’ Institution. There were no constraints placed on the schedule based on staffing or patient beds in the recovery unit or ward.

#### 2.3.2 Optimization formulations

Three scheduling optimization problems were formulated. All formulations were based on an integer linear programming framework that has been used for many different scheduling problems including but not limited to surgery scheduling ^24,30^. The first, “Any”, was adapted from Marques et al. ^24^ with notable modifications including the addition of an overtime penalty and only considering one surgery specialty. “Any” allows any surgery to be scheduled at any time in the day in any room subject to the constraints that no surgeries in a room overlap, and that no surgeon is operating in two rooms simultaneously. The second formulation, “Split”, is the same as “Any” but enforces a maximum of 2 surgeons per OR on a given day and a maximum of 1 room per surgeon per day. Finally, the third formulation is akin to a max-sum multiple subset sum problem and is thus referred to as “MSSP”. It enforces 1 surgeon per OR on a given day, simplified into multiple optimization problems for each surgeon, following a fair distribution of rooms among surgeons. “Any” is the most flexible optimization formulation, i.e., the least constrained of the three. “Split” and “Any” impose additional constraints on feasible schedules and reflect realistic logistical restrictions that a hospital may want to impose, e.g., having a surgeon use the same room on any given day. Additional benefits to these more constrained formulations will be explored further in the paper.

#### 2.3.3 Notation

We use *C* for the set of all surgeries under consideration, *R* for the set of all operating rooms, and *D* for the set of all days in the planning horizon. Time is discretized into slots with granularity *g* (e.g., 10 minutes or 15 minutes). *T*_*c*_ is the set of all time slots in a day and *T*_*o*_ is the subset of *T*_*c*_ consisting of overtime slots (e.g., after 5pm), for which a penalty will be applied. *p*_*c*_ is the duration of surgery for surgery *c* divided by granularity *g. γ* is the time needed for cleaning the rooms between operations divided by granularity *g. λ* is the weight for the penalty of going overtime. *C*_2_ is the set of surgeries that have high priority, i.e., that must be scheduled mandatorily in the planning horizon. The subscript of 2 in *C*_2_ is used to be consistent with past work done to define priority groups ^24^. *H* is the set of all surgeons, and *h*_*c*_ is the surgeon that is assigned to surgery *c*. Binary indicators *i*_*hd*_ are used to denote whether surgeon *h* is available on day *d*.

We are now ready to describe the three optimization formulations mathematically.

#### 2.3.4 The “Any” formulation

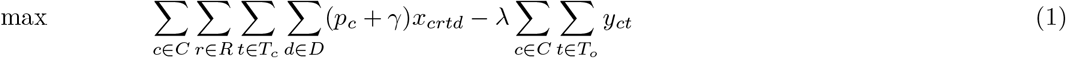

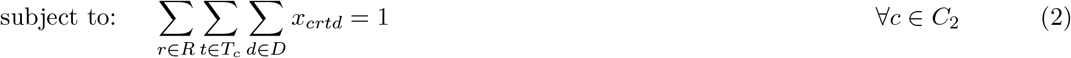

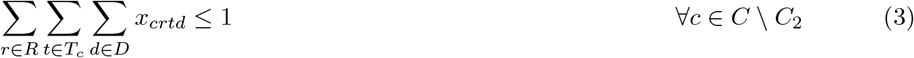

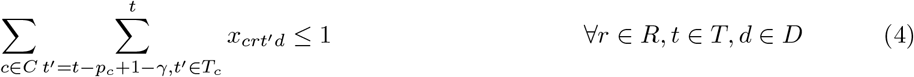

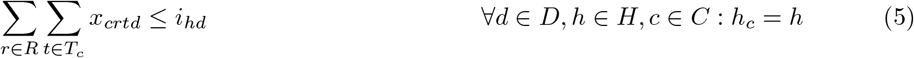

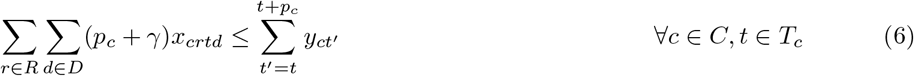

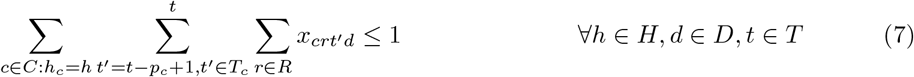

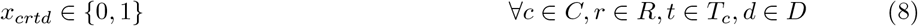

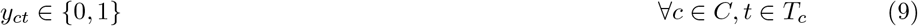

This is a scheduling problem with both soft and hard time constraints. The soft constraint allows surgeries to be scheduled over OR closing time, incurring an overtime penalty. However, there is also a hard constraint of 1 hour after closing that is reflected in *T*_*c*_. The objective (1) is to maximize the total OR utliization time (first term) while penalizing any overtime using a penalty term, *λ* (second term). The decision variables defined in Constraints (8, 9) are *x*_*crtd*_ and *y*_*ct*_, all of which are binary, where *x*_*crtd*_ is 1 if surgical case *c* is assigned to room *r* and performed starting at time slot *t* on day *d*, and *y*_*ct*_ is 1 if surgery *c* is performed in time slot *t*. Constraint (2) ensures that patients with high priority are scheduled within the planning horizon. Constraint (3) enforces that a surgery can only be done once. Constraint (4) ensures that there is no overlap between surgeries in a room, including a cleaning time of *γ*. Constraint (5) ensures that a surgery cannot be scheduled if the surgeon is unavailable on that day. Constraint (6) defines the relation between *x* and *y*. Constraint (7) ensures that there is no overlap between surgeries performed by the same surgeon across different rooms.

#### 2.3.5 The “Split” formulation

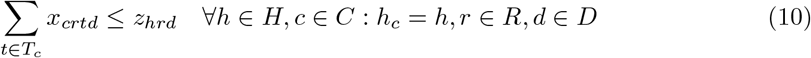

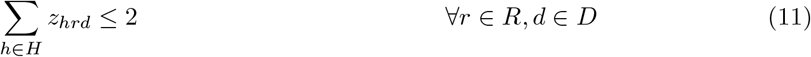

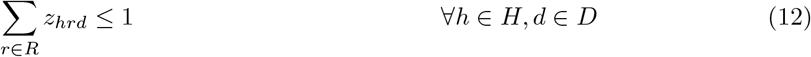

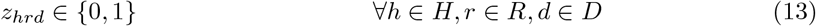

“Split” is almost identical to the Any formulation, with a few modifications. A new binary variable, *z*_*hrd*_, is introduced in Constraint (13) to define whether a surgeon *h* is performing any surgeries in room *r* on day *d*. Constraint (10) defines the relation between *x* and *z*. Constraint (11) ensures that there are at most 2 surgeons performing surgery in any one operating room, while Constraint (12) ensures that a surgeon only performs surgery in one room on a given day.

#### 2.3.6 The Max-Sum Multiple Subset Sum Problem (“MSSP”) formulation

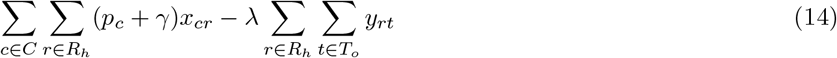

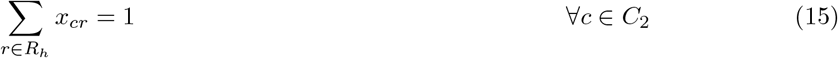

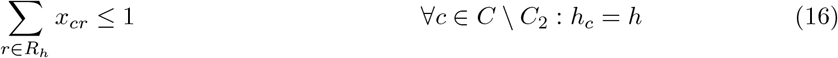

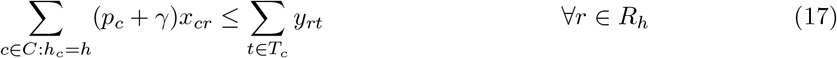

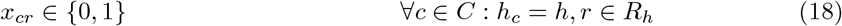

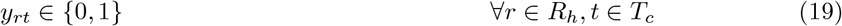

The MSSP optimization problem was solved for each surgeon’s wait list, and then the results of each optimization problem were combined into one schedule. The solution is independent of the order in which each subpart of the MSSP problem was solved, as the operating room assignments are done prior to the solve. Operating rooms were assigned to each surgeon by giving each surgeon an equal number of rooms, with any surplus rooms assigned to the surgeons with the most availability.

The two sets of binary decision variables are *x*_*cr*_, which is set to 1 if a surgery *c* is performed in room *r*, and *y*_*rt*_, which is set to 1 if a surgery is being performed in room *r* at time slot *t*. In this formulation, *R*_*h*_ refers to the rooms that are assigned in advance to each surgeon; each room is used on a different day. The objective (14) is similar to the one from “Any” and “Split”. Constraint (15) ensures that the priority cases are done in the planning horizon, and Constraint (16) ensures that a surgery is performed at most once. Constraint (17) defines the relationship between *x* and *y*.

### 2.4 Simulated Schedule Generation

Using the ACS NSQIP data, simulated schedules were generated using each optimization formulation, with schedule parameters as follows: Schedule granularity of 10- and 15-minute block times were considered. These sizes were chosen to ensure interpretable schedule generation. Surgery completion times were rounded up to the nearest block. The effect of surgeon wait list size on schedule accuracy was also evaluated. Wait list sizes representing 2 weeks (*n* = 250), 4 weeks (*n* = 500), 8 weeks (*n* = 1000) and 12 weeks (*n* = 1500) were considered. The cases within the wait list were all given the same priority to be booked (no rank by time), unless randomly considered to be of high priority.

Three different versions of each model with the above schedule parameters were created. The first model used the ML-predicted DOS values for THA and TKA; this is the novel two-stage approach of predicting then optimizing that we are proposing in this work. The second model used the mean DOS for each different type of surgery (THA and TKA) in order to obtain a schedule that mimics the current OR schedule. Finally, a hindsight model was created that used the true DOS values in order to provide an upper bound on the best possible schedule that could be generated with perfect information (100% accurate to the minute). Each of these 3 simulations was performed 104 times (to represent 2 years) using a random sample of surgeries from the testing set. Although the case mix was not fixed, the testing set had an approximately even number of THAs compared to TKAs.

### 2.5 Scheduling Comparisons

To compare the three schedule optimization formulations (Any, Split, MSSP), the results of the two-stage predict-then-optimize simulation results across all schedule parameter combinations were assessed. To evaluate the efficacy of the two-stage schedule generation technique, it was compared to the results of scheduling by two other techniques; the mean DOS for each surgery (the current gold-standard at most institutions) and the hindsight schedule. Each of these three schedules was constructed for each week of the simulation, and metrics consisting of overtime, underutilization, and the objective function value were calculated for each schedule. Student’s t-test was used to compare the effect between scheduling formulations. Overtime and underutilization for two-stage and mean DOS schedules over the simulated weeks were compared using the paired Wilcoxon signed-rank test as the same surgical cases were randomly selected for each generated schedule. The effect of schedule granularity and considered wait list size were evaluated using unpaired Student’s t-test and analysis of variance as these were grouped over multiple selections of random cases. P-values of *<*0.05 were considered statistically significant.

## 3 Results

### 3.1 Prediction Model Accuracy

The TKA prediction model obtained a training accuracy (with a 30 minute buffer) of 76.9% and a training mean squared error (MSE) of 0.904. The validation accuracy was 77.7% with an MSE of 0.904. The test accuracy was 78.1% with an MSE of 0.898. The THA prediction model obtained a training accuracy of 74.0% and a training MSE of 0.888. The validation accuracy was 75.0% with an MSE of 0.910, and the test accuracy was 75.4% with an MSE of 0.916.

### 3.2 Schedule Optimization Formulation Comparison

Overall, the Any scheduling optimization formulation exhibited the poorest performance across all different schedule parameter combinations (Table 1). There was no significant difference in overtime across all schedule parameters between the Split and MSSP formulations. For two combinations of schedule parameters (10-minute granularity, 1500 wait list size and 15-minute granularity, 1500 wait list size), there was significantly less OR underutilization with the MSSP formulation, however, this was only 10.9 minutes and 15.4 minutes, respectively, on average over an entire week (Table 1). In contrast, for 15-minute granularity and a 250 wait list size, there was significantly more OR underutilization with MSSP compared to the Split formulation, 42.3 minutes over an entire week (Table 1). Figure 2 and Figure 3 display this comparison for overtime and underutilization across all three optimization problems, respectively.

**Table 1.**
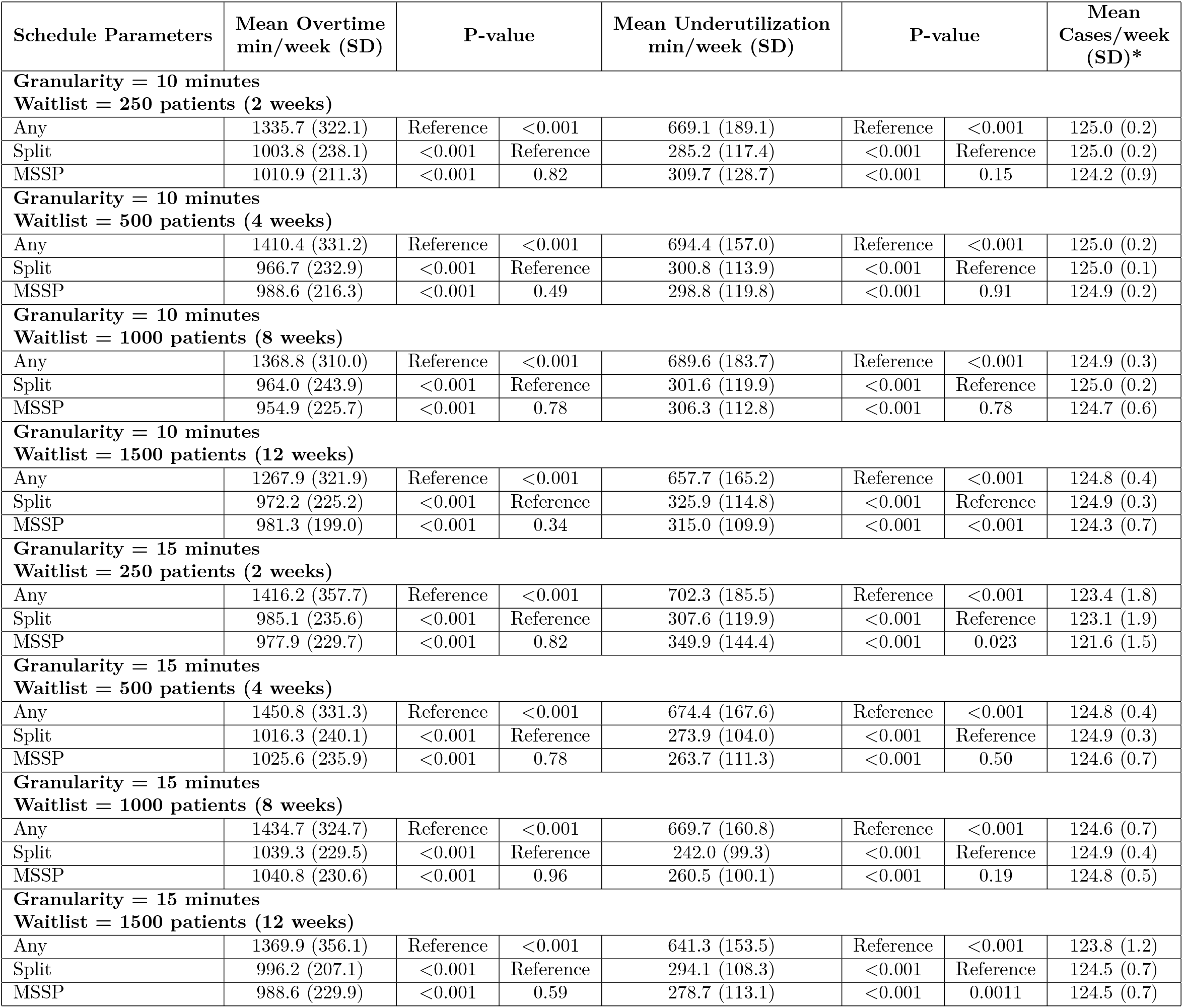
Simulated schedule results for two-stage (predict-then-optimize) for each schedule optimization formulation. *Mean number of cases/week not significant between any schedule optimization formulations.

**Figure 2.**
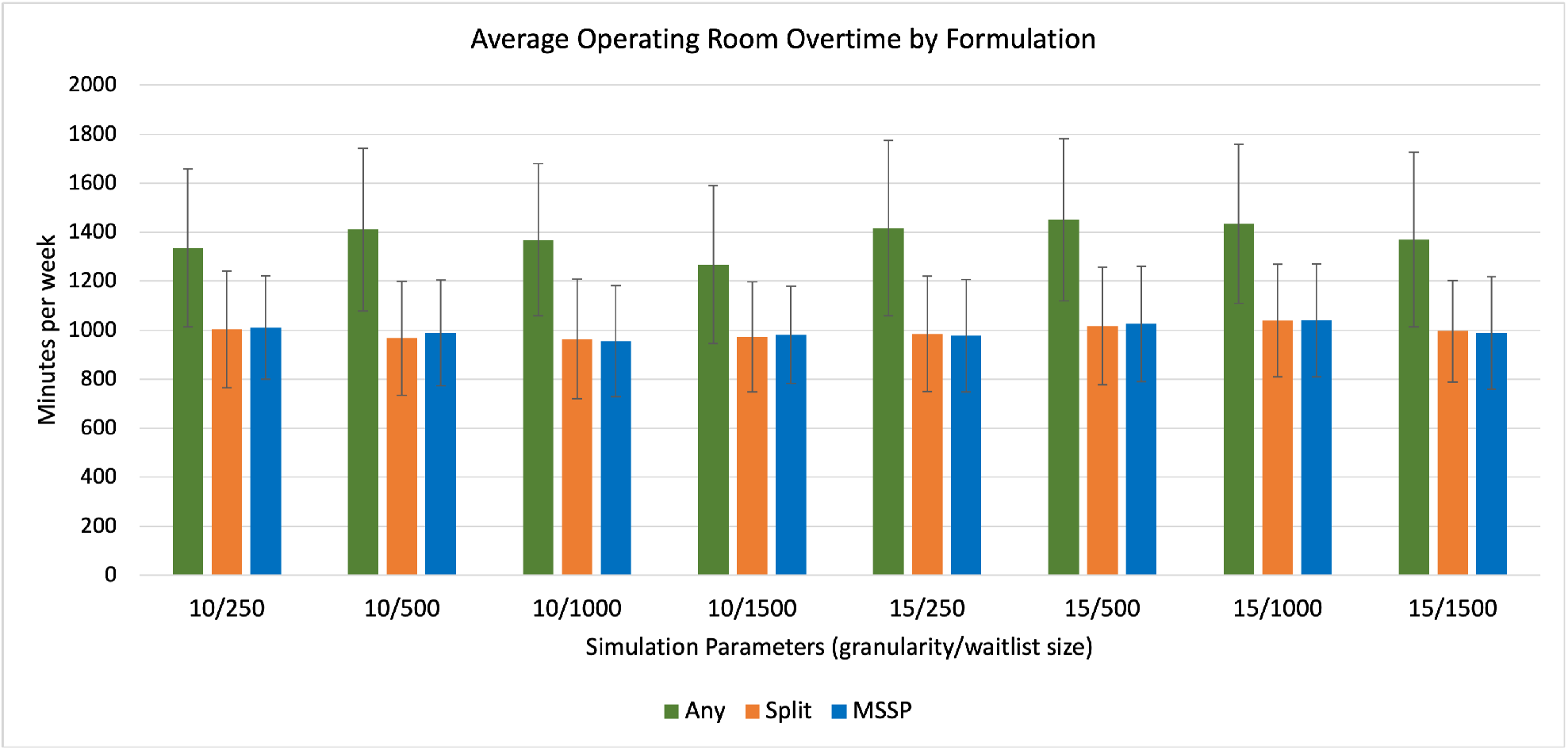
Mean overtime for each schedule optimization formulation across all schedule parameter combinations. MSSP = Multiple Subset Sum Problem

**Figure 3.**
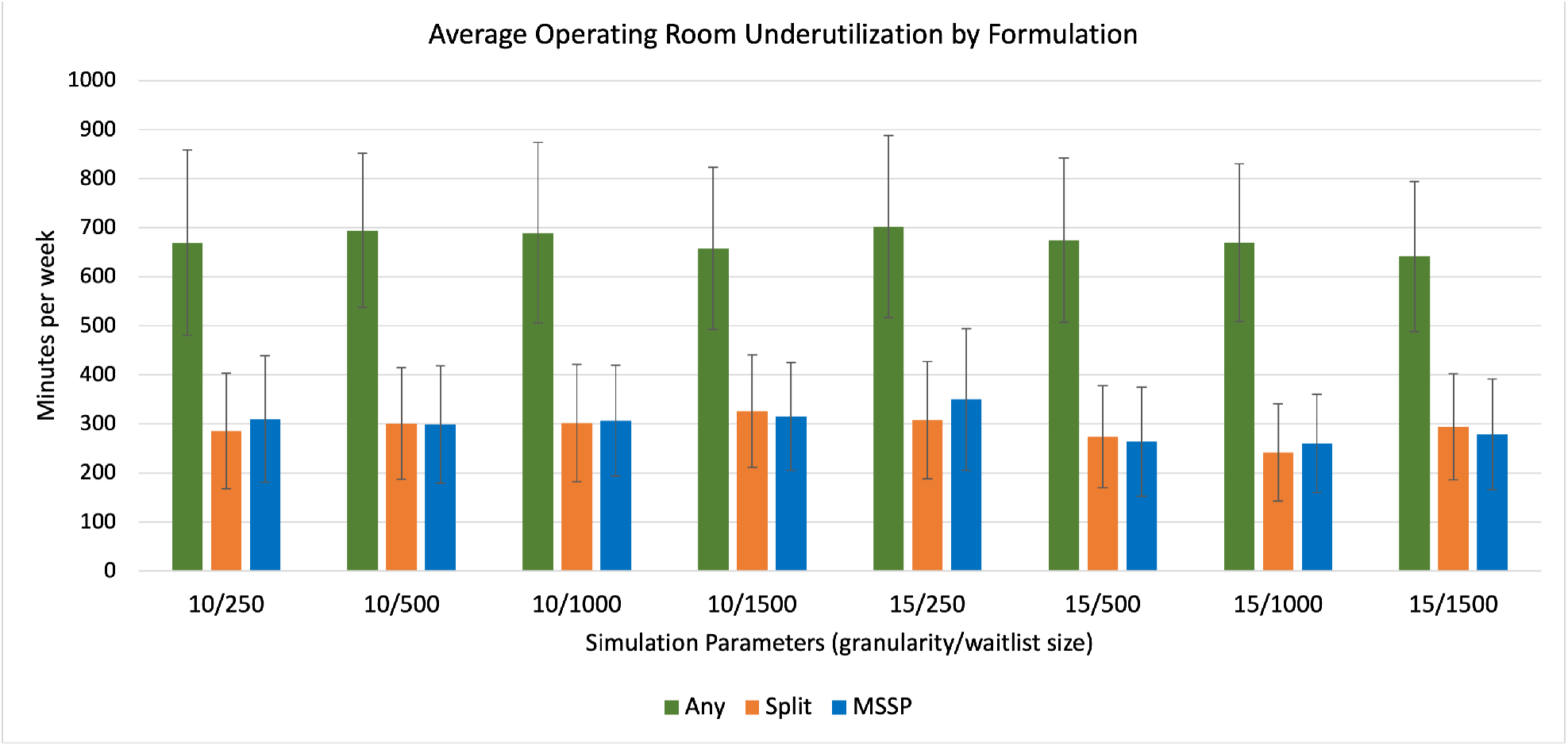
Mean underutilization for each schedule optimization formulation across all schedule parameter combinations. MSSP = Multiple Subset Sum Problem

### 3.3 Simulated Schedule Comparison

As the MSSP scheduling formulation performed best across all schedule parameters, it was used in all further analyses. The two-stage predict-then-optimize approach performed better than using mean DOS for over 80% of weekly schedules in terms of objective optimization problem value across all schedule parameter combinations. This difference was more consistent across the schedules generated using 15-minute schedule granularity size, where two-stage was superior to mean in over 90% of simulated schedules (Table 2). There was less overtime across all schedule parameters when utilizing the two-stage approach (p*<*0.001), equating to an average decrease in overtime of 300 to 500 minutes per week at the simulated hospital (or 12 to 20 minutes per operating room per day). However, there was more OR underutilization with the two-stage approach across all schedule parameters (Table 3, p*<*0.001). As expected, the hindsight schedule was nearly perfect for all generated schedules and was significantly better than the two-stage or mean approach with respect to objective value, overtime and underutilization (Table 3). Despite a statistically significant difference, there was no clinically realizable difference in the number of cases performed between two-stage and mean groups, however, the hindsight formulation scheduled less cases than both the mean and the two-stage approach (mean of approximately 12 fewer cases per week).

**Table 2.**
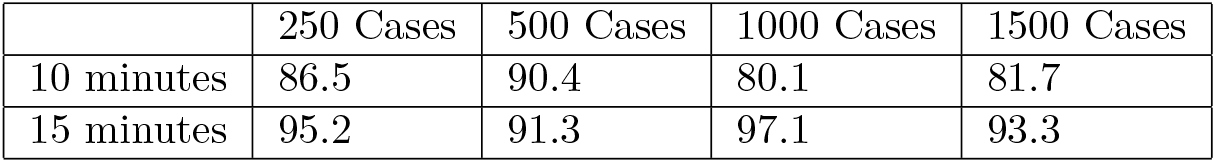
Percentage of simulations in which the two-stage performed better than mean for all schedule parameters using the MSSP schedule optimization formulation.

**Table 3.**
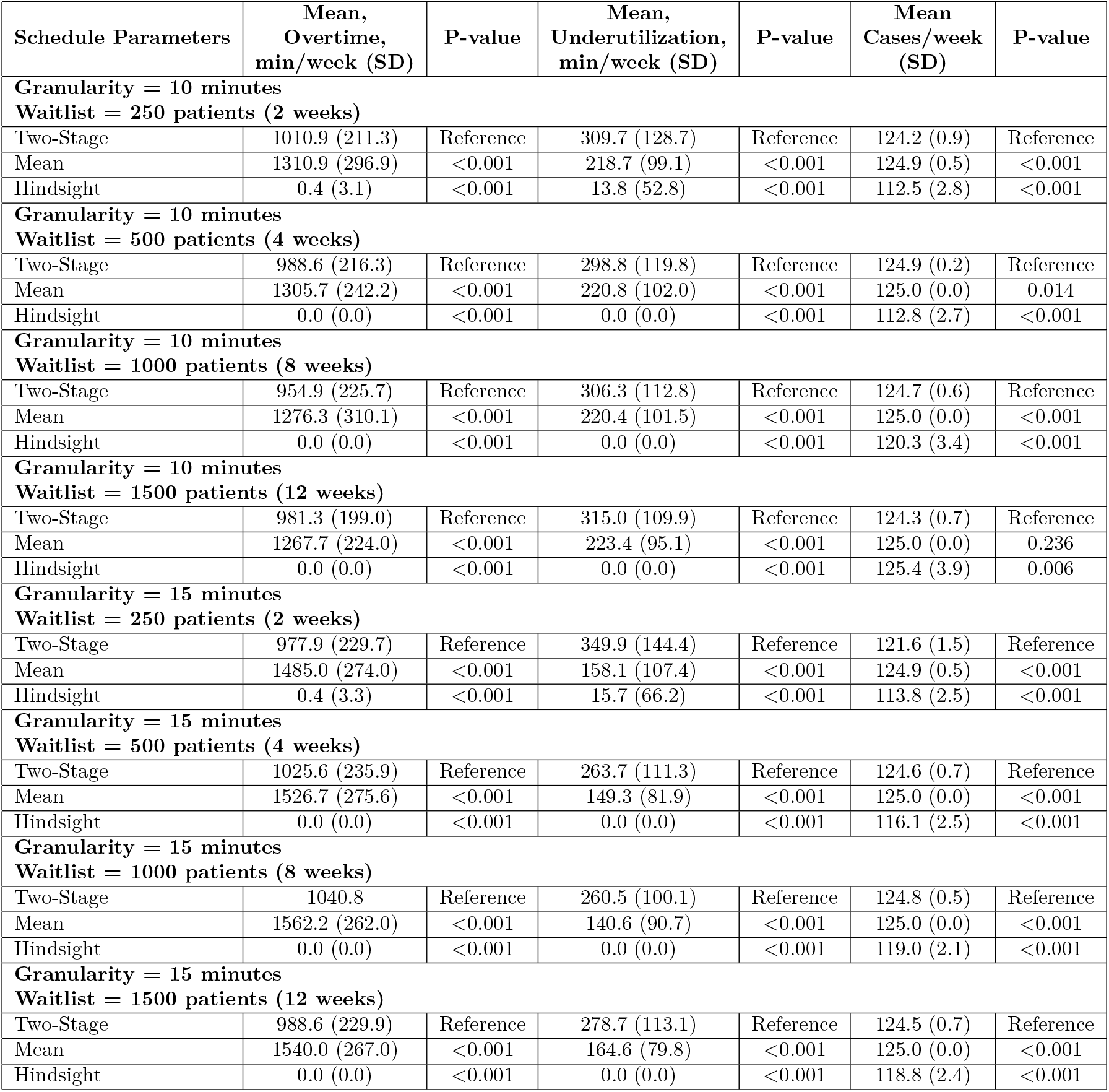
Comparing two-stage, mean and hindsight duration of surgery using the Multiple Subset Sum Problem (MSSP) schedule optimization formulation. SD = Standard deviation

The changes to schedule granularity and considered wait list size did not influence the amount of overtime. However, there was significantly less OR underutilization with the 15-minute granularity schedules (p=0.022) and with wait list sizes greater than 500, or 1 month considered (p*<*0.001) (Table 4).

**Table 4.**
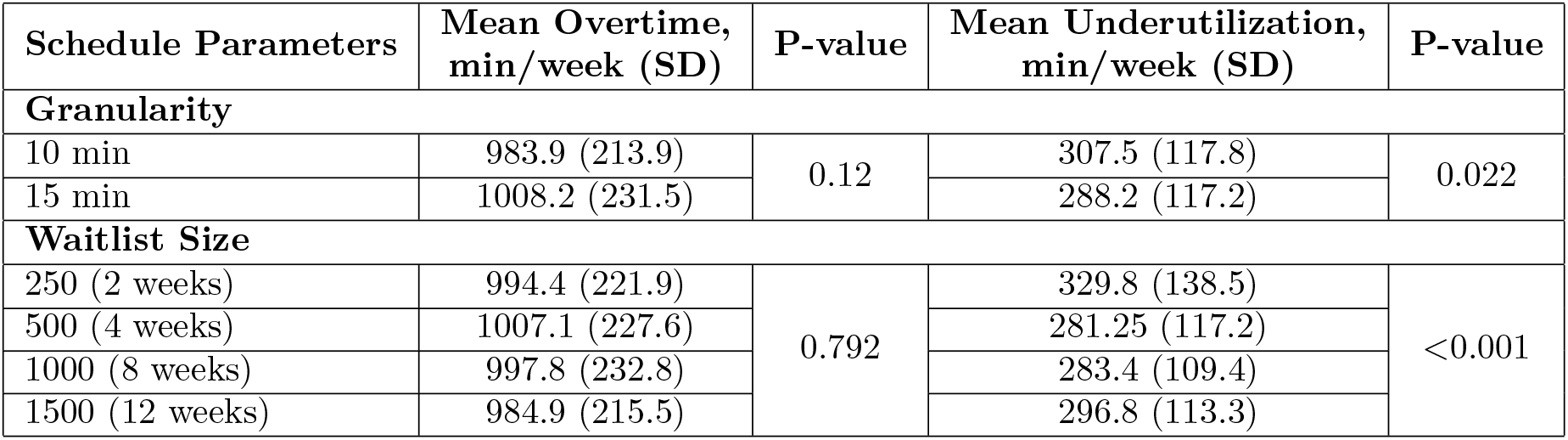
Comparing impact of schedule parameters for two-stage Multiple Subset Sum Problem (MSSP). SD = Standard deviation

## 4 Conclusion and Discussion

This paper compared three different scheduling optimization problems and evaluated a novel approach to surgical scheduling for THA and TKA utilizing a combined two-stage ML DOS prediction then optimization. There was no significant difference in OR underutilization or overtime between the MSSP (one surgeon designated to one OR per day) or Split (maximum of two surgeons designated for one OR per day) optimization formulations. However, these both performed significantly better than the Any (no limit on surgeons per OR per day) formulation. We believe this is due to the limitations of the predictions: underestimating the DOS of one case *c* being performed by surgeon *h* in room *r* can have cascading effects on another room *r*^*′*^ in which the same surgeon *h* is due to perform another surgery *c*^*′*^ at a later time. This causes additional overtime penalties for “Any”, something that the more restrictive “Split” and “MSSP” do not encounter. This is why despite optimal solutions to “Any” being theoretically better than those of “Split”, they performed worse when simulated with the actual DOS.

Overall, the combined two-stage approach significantly outperformed the current standard for scheduling cases which is a case-specific mean surgery duration. This performance improvement was maintained across all schedule parameter combinations, including different schedule block granularity and different patient wait list sizes considered. Despite this improvement, the two-stage approach performed considerably worse than the hindsight schedule, highlighting the limitations of the current predictions that are based solely on preoperative patient data. Interestingly, there was no impact on overtime by varying schedule granularity or wait list size, however, both of these impacted the amount of underutilization. The smaller wait list size of two weeks of considered cases had a greater amount of underutilization, which was likely due to surgeons not having enough cases to fill their OR time. Once a threshold was met at a four week pool of cases there was no difference between groups. Also notable was the fact that when only considering a two week pool of cases, the Split optimization formulation outperformed MSSP, likely due to some surgeons not having enough cases to fill their time. Therefore, as the MSSP formulation is most practically implementable, it must be ensured that either the considered case pool is large enough or surgeons are allocated time when they have enough cases to fill an entire OR day to avoid underutilization.

Errors in procedure length estimation by clinicians occur in approximately 75% of cases, with 32-50% of daily OR schedules being underbooked and 37-42% overbooked ^31,32^. This is compounded by the fact that less than 50% of operating time is actually spent doing surgery ^31^. Booking based on a historical mean is more accurate than when estimated by the surgical team, though less accurate than traditional ML approaches ^10,33–35^. Previous approaches using computing to improve surgical scheduling have included schedule optimization or ML to predict DOS in isolation ^13,19,24,35–37^. Other approaches to schedule OR utilization include the of use surgeon-specific mean DOS or a surgeon case-specific rolling average time. To our knowledge, these have not been compared to an ML prediction-based approach. Due to the lack of surgeon-specific details included in the ACS NSQIP dataset, we could not assess the efficacy of these approaches in the current study.

The implementation of this predict-then-optimize scheduling approach would face several challenges in the real world. The MSSP optimization model is in line with current surgical scheduling practices at most hospitals. This formulation optimizes a specific surgeon’s waiting list, increasing their ability to accurately plan their day while ensuring a fair distribution of time (by OR days) for each surgeon. However, attempting to implement the other optimization formulations (Split and Any) may be faced with resistance by end-users. Particularly, using the Any formulation, surgeons may have cases at the beginning and end of the day spread out across more days in a week. ML-predicted DOS has been trialed previously in OR planning by one group that found a reduction in wait time between cases ^38^. However, they generated the predicted DOS and evaluated the implications of that information over a single day, not considering other cases from the wait list or optimizing the schedule based on the predicted DOS.

The generated models and optimization formulations have the ability to transform how elective OR scheduling is performed. By developing models specific to each operation, this increases the accuracy of each model. Most previous research evaluating the effect of ML for DOS prediction has grouped multiple different procedures ^19,35^. Using such models, the procedure performed would generally be the most important feature, diluting the effect of other important patient factors without using appropriate ML techniques. The potential for cost savings for hospitals, related to reduction in overtime costs and valuable underutilized OR time, are high, but the main limitation lies in the accuracy of the DOS prediction. The predictive models for TKA and THA included 33 individual patient features. Improving the model with the use of operational factors from a specific institution would likely have a corresponding effect on the schedule results but would reduce the generalizability of the approach. Further improvements may also be made by directly integrating the downstream scheduling optimization problem into training the predictive ML model ^39^.

In this work the DOS prediction was restricted to preoperative patient factors, based on the availability of data elements in the ACS NSQIP database, limiting the prediction accuracy. This was evidenced by the large difference in schedule performance between the two-stage method and the perfect, hindsight, schedule. Nevertheless, this approach to predicting DOS still yielded improved schedules as compared to utilizing a surgery-specific mean time estimate. Secondly, the goal of our optimization problem was to maximize the utilization of the OR, however, this may not be directly in line with the goals of all hospitals, as some institutions may have other priorities, such as maximizing the number of cases completed. This project only developed predictive models for primary TKA and THA procedures which may have reduced the artificially lowered the potential effect size of using this approach as these are relatively routine procedures with lower DOS variability. By generating more surgery-specific predictive models within orthopaedic surgery, or other specialties, the potential for this approach may be even larger. However, these results are more directly applicable to high-volume arthroplasty surgical centres. Additionally, this scheduling approach did not consider downstream constraints, such as number of recovery beds, ward beds or available staff. Finally, the Split and Any optimization problems were computationally intensive when considering a waiting list size of 2-3 months of patients. This may be a consideration depending on institutional computational resources, which is worth noting when implementing a similar solution.

### 4.2 Conclusion

Planning operating room schedules by individual surgeons or splitting them by a maximum of two surgeons provided comparable schedules over two years of simulated schedule generation. Using ML patient-specific DOS predictions coupled with optimization, was superior to elective scheduling based on a mean DOS metric over three different optimization problems with varying constraints, combinations of wait list size and granularity. This generalizable approach suggests that improvements in hospital resource utilization are possible with application of new computational methods, but inclusion of institution-specific operational data may be considered to further improve predictions and scheduling. This has significant potential implications for healthcare systems struggling with pressures of rising costs and growing operative wait lists.

## Data Availability

All data produced in the present study are available upon request and approval from the American College of Surgeons.

https://www.facs.org/quality-programs/data-and-registries/acs-nsqip/

## Acknowledgements

Database: The American College of Surgeons National Surgical Quality Improvement Program and the hospitals participating in the ACS NSQIP are the source of the data used herein; they have not verified and are not responsible for the statistical validity of the data analysis or the conclusions derived by the authors.

## Appendix

***Appendix A: Supplementary Table 1***

**Supplementary Table 1.** Comparing two-stage, mean, and hindsight duration of surgery using the Any schedule optimization formulation.

| Schedule Parameters | Mean,<br>Overtime,<br>min/week (SD) | P-value | Mean,<br>Underutilization,<br>min/week (SD) | P-value | Mean<br>Cases/week<br>(SD) | P-value |
| --- | --- | --- | --- | --- | --- | --- |
| **Granularity = 10 minutes** |  |  |  |  |  |  |
| **Waitlist = 250 patients (2 weeks)** |  |  |  |  |  |  |
| Two-Stage | 1335.7 (322.1) | Reference | 669.1 (189.1) | Reference | 125.0 (0.2) | Reference |
| Mean | 1706.5 (372.1) | <0.001 | 655.7 (177.7) | 0.948 | 125.0 (0.0) | 0.083 |
| Hindsight | 0.0 (0.0) | <0.001 | 0.0 (0.0) | <0.001 | 117.2 (2.8) | <0.001 |
| **Granularity = 10 minutes** |  |  |  |  |  |  |
| **Waitlist = 500 patients (4 weeks)** |  |  |  |  |  |  |
| Two-Stage | 1410.4 (331.2) | Reference | 694.4 (157.0) | Reference | 125.0 (0.2) | Reference |
| Mean | 1728.1 (354.8) | <0.001 | 644.0 (178.4) | 0.011 | 125.0 (0.0) | 0.025 |
| Hindsight | 0.0 (0.0) | <0.001 | 0.0 (0.0) | <0.001 | 121.0 (4.5) | <0.001 |
| **Granularity = 10 minutes** |  |  |  |  |  |  |
| **Waitlist = 1000 patients (8 weeks)** |  |  |  |  |  |  |
| Two-Stage | 1368.8 (310.0) | Reference | 689.6 (183.7) | Reference | 126.9 (0.3) | Reference |
| Mean | 1804.0 (366.8) | <0.001 | 698.5 (180.9) | 0.745 | 126.9 (0.4) | 0.564 |
| Hindsight | 0.0 (0.0) | <0.001 | 0.8 (7.8) | <0.001 | 126.4 (5.8) | 0.764 |
| **Granularity = 10 minutes** |  |  |  |  |  |  |
| **Waitlist = 1500 patients (12 weeks)** |  |  |  |  |  |  |
| Two-Stage | 1267.9 (321.9) | Reference | 657.7 (165.2) | Reference | 126.8 (0.4) | Reference |
| Mean | 1452.4 (378.0) | <0.001 | 883.2 (287.2) | <0.001 | 123.0 (3.3) | <0.001 |
| Hindsight | 0.0 (0.0) | <0.001 | 0.0 (0.0) | <0.001 | 130.6 (4.1) | <0.001 |
| **Granularity = 15 minutes** |  |  |  |  |  |  |
| **Waitlist = 250 patients (2 weeks)** |  |  |  |  |  |  |
| Two-Stage | 1416.2 (357.7) | Reference | 702.3 (185.5) | Reference | 123.4 (1.8) | Reference |
| Mean | 2001.3 (366.2) | <0.001 | 622.9 (176.1) | 0.007 | 125.0 (0.0) | <0.001 |
| Hindsight | 0.0 (0.0) | <0.001 | 0.0 (0.0) | <0.001 | 117.3 (2.7) | <0.001 |
| **Granularity = 15 minutes** |  |  |  |  |  |  |
| **Waitlist = 500 patients (4 weeks)** |  |  |  |  |  |  |
| Two-Stage | 1450.8 (331.3) | Reference | 674.4 (167.6) | Reference | 124.8 (0.4) | Reference |
| Mean | 2048.7 (364.7) | <0.001 | 661.6 (171.9) | 0.304 | 125.0 (0.0) | <0.001 |
| Hindsight | 0.0 (0.0) | <0.001 | 0.0 (0.0) | <0.001 | 121.0 (2.7) | <0.001 |
| **Granularity = 15 minutes** |  |  |  |  |  |  |
| **Waitlist = 1000 patients (8 weeks)** |  |  |  |  |  |  |
| Two-Stage | 1434.7 (324.7) | Reference | 669.8 (160.8) | Reference | 124.6 (0.7) | Reference |
| Mean | 2077.2 (396.5) | <0.001 | 663.6 (198.8) | 0.843 | 125.0 (0.0) | <0.001 |
| Hindsight | 0.0 (0.0) | <0.001 | 0.0 (0.0) | <0.001 | 122.6 (3.3) | <0.001 |
| **Granularity = 15 minutes** |  |  |  |  |  |  |
| **Waitlist = 1500 patients (12 weeks)** |  |  |  |  |  |  |
| Two-Stage | 1369.9 (356.1) | Reference | 641.3 (153.5) | Reference | 123.8 (1.2) | Reference |
| Mean | 2008.6 (370.0) | <0.001 | 599.4 (162.5) | 0.028 | 125.0 (0.0) | <0.001 |
| Hindsight | 0.0 (0.0) | <0.001 | 0.0 (0.0) | <0.001 | 125.2 (5.6) | 0.006 |

***Appendix B: Supplementary Table 2***

**Supplementary Table 2.** Comparing two-stage, mean, and hindsight duration of surgery using the Split schedule optimization formulation.

| Schedule Parameters | Mean,<br>Overtime,<br>min/week (SD) | P-value | Mean,<br>Underutilization,<br>min/week (SD) | P-value | Mean<br>Cases/week<br>(SD) | P-value |
| --- | --- | --- | --- | --- | --- | --- |
| **Granularity = 10 minutes** |  |  |  |  |  |  |
| **Waitlist = 250 patients (2 weeks)** |  |  |  |  |  |  |
| Two-Stage | 1003.8 (238.1) | Reference | 285.2 (117.4) | Reference | 124.9 (0.2) | Reference |
| Mean | 1356.0 (277.4) | <0.001 | 194.9 (94.1) | <0.001 | 125.0 (0.0) | 0.046 |
| Hindsight | 0.0 (0.0) | <0.001 | 0.0 (0.0) | <0.001 | 116.1 (3.0) | <0.001 |
| **Granularity = 10 minutes** |  |  |  |  |  |  |
| **Waitlist = 500 patients (4 weeks)** |  |  |  |  |  |  |
| Two-Stage | 966.7 (232.9) | Reference | 300.8 (113.9) | Reference | 125.0 (0.1) | Reference |
| Mean | 1253.3 (277.9) | <0.001 | 212.5 (92.0) | <0.001 | 125.0 (0.0) | 0.317 |
| Hindsight | 0.0 (0.0) | <0.001 | 0.0 (0.0) | <0.001 | 120.5 (3.3) | <0.001 |
| **Granularity = 10 minutes** |  |  |  |  |  |  |
| **Waitlist = 1000 patients (8 weeks)** |  |  |  |  |  |  |
| Two-Stage | 964.0 (243.9) | Reference | 301.6 (119.9) | Reference | 125.0 (0.2) | Reference |
| Mean | 1319.7 (289.7) | <0.001 | 216.9 (106.8) | <0.001 | 125.0 (0.0) | 0.025 |
| Hindsight | 0.9 (8.8) | <0.001 | 5.4 (55.6) | <0.001 | 119.9 (3.9) | <0.001 |
| **Granularity = 10 minutes** |  |  |  |  |  |  |
| **Waitlist = 1500 patients (12 weeks)** |  |  |  |  |  |  |
| Two-Stage | 972.2 (225.2) | Reference | 325.9 (114.8) | Reference | 124.9 (0.3) | Reference |
| Mean | 1258.9 (265.6) | <0.001 | 335.1 (241.6) | 0.497 | 123.8 (2.4) | <0.001 |
| Hindsight | 12.8 (20.7) | <0.001 | 176.6 (248.5) | <0.001 | 116.3 (5.3) | <0.001 |
| **Granularity = 15 minutes** |  |  |  |  |  |  |
| **Waitlist = 250 patients (2 weeks)** |  |  |  |  |  |  |
| Two-Stage | 985.1 (235.6) | Reference | 307.6 (119.9) | Reference | 123.1 (1.9) | Reference |
| Mean | 1497.3 (308.5) | <0.001 | 156.3 (78.7) | <0.001 | 125.0 (0.0) | <0.001 |
| Hindsight | 0.0 (0.0) | <0.001 | 0.0 (0.0) | <0.001 | 114.5 (2.9) | <0.001 |
| **Granularity = 15 minutes** |  |  |  |  |  |  |
| **Waitlist = 500 patients (4 weeks)** |  |  |  |  |  |  |
| Two-Stage | 1016.3 (240.1) | Reference | 273.9 (104.0) | Reference | 124.9 (0.3) | Reference |
| Mean | 1516.7 (228.9) | <0.001 | 153.9 (84.1) | <0.001 | 125.0 (0.0) | <0.001 |
| Hindsight | 0.0 (0.0) | <0.001 | 0.0 (0.0) | <0.001 | 116.6 (5.4) | <0.001 |
| **Granularity = 15 minutes** |  |  |  |  |  |  |
| **Waitlist = 1000 patients (8 weeks)** |  |  |  |  |  |  |
| Two-Stage | 1039.3 (229.5) | Reference | 242.0 (99.3) | Reference | 124.9 (0.4) | Reference |
| Mean | 1538.8 (272.9) | <0.001 | 139.0 (71.2) | <0.001 | 125.0 (0.0) | <0.001 |
| Hindsight | 0.0 (0.0) | <0.001 | 0.0 (0.0) | <0.001 | 117.8 (5.4) | <0.001 |
| **Granularity = 15 minutes** |  |  |  |  |  |  |
| **Waitlist = 1500 patients (12 weeks)** |  |  |  |  |  |  |
| Two-Stage | 996.2 (207.1) | Reference | 294.1 (108.3) | Reference | 124.5 (0.7) | Reference |
| Mean | 1531.9 (245.3) | <0.001 | 149.9 (71.9) | <0.001 | 125.0 (0.0) | <0.001 |
| Hindsight | 3.6 (36.6) | <0.001 | 0.0 (0.0) | <0.001 | 120.5 (4.3) | <0.001 |

## References

1. Healthcare cost and Utilization Project (HCUP); 2021. Available from: https://www.ahrq.gov/data/hcup/index.html.

2. Demography - elderly population - OECD data; 2022. Available from: https://data.oecd.org/pop/elderly-population.htm.

3. The Lancet Gastroenterology & Hepatology. Obesity: another ongoing pandemic. The Lancet Gastroenterology & Hepatology. 2021;6(6):411. Available from: https://www.sciencedirect.com/science/article/pii/S2468125321001436.

4. Muñoz E, Muñoz W, Wise L. National and Surgical Health Care Expenditures, 2005-2025. Annals of surgery. 2010 02;251:195–200.

5. BMUS: The burden of musculoskeletal diseases in the United States; 2011. Available from: https://www.boneandjointburden.org/.

6. Scott CEH, MacDonald DJ, Howie CR. ‘Worse than death’ and waiting for a joint arthroplasty. The Bone & Joint Journal. 2019;101-B(8):941–50. PMID: 31362549. Available from: 10.1302/0301-620X.101B8.BJJ-2019-0116.R1.

7. Viberg N, Forsberg BC, Borowitz M, Molin R. International comparisons of waiting times in health care – Limitations and prospects. Health Policy. 2013;112(1):53–61. Available from: https://EconPapers.repec.org/RePEc:eee:hepoli:v:112:y:2013:i:1:p:53-61.

8. Rozell JC, Ast MP, Jiranek WA, Kim RH, Della Valle CJ. Outpatient Total Joint Arthroplasty: The New Reality. J Arthroplasty. 2021 Feb;36(7S):S33–9.

9. Attarian DE, Wahl JE, Wellman SS, Bolognesi MP. Developing a high-efficiency operating room for total joint arthroplasty in an academic setting. Clin Orthop Relat Res. 2013 Jun;471(6):1832–6.

10. Huang CC, Lai J, Cho DY, Yu J. A Machine Learning Study to Improve Surgical Case Duration Prediction. medRxiv. 2020. Available from: https://www.medrxiv.org/content/early/2020/06/12/2020.06.10.20127910.

11. Bartek MA, Saxena RC, Solomon S, Fong CT, Behara LD, Venigandla R, et al. Improving Operating Room Efficiency: Machine Learning Approach to Predict Case-Time Duration. J Am Coll Surg. 2019 Jul;229(4):346-54.e3.

12. Martinez O, Martinez C, Parra CA, Rugeles S, Suarez DR. Machine learning for surgical time prediction. Comput Methods Programs Biomed. 2021 Jun;208:106220.

13. Marques I, Captivo M, Barros N. Optimizing the master surgery schedule in a private hospital. Operations Research for Health Care. 2018 12;20.

14. Erdogan SA, Denton BT. In: Surgery Planning and Scheduling. John Wiley & Sons, Ltd; 2011. Available from: https://onlinelibrary.wiley.com/doi/abs/10.1002/9780470400531.eorms0861.

15. Shi Y, Mahdian S, Blanchet J, Glynn P, Shin AY, Scheinker D. Surgical Scheduling via Optimization and Machine Learning with Long-Tailed Data. Health Care Management Science. 2022 09;In Press:In Press.

16. Ahmed A, He L, Chou Ca, Hamasha MM. A prediction-optimization approach to surgery prioritization in operating room scheduling. Journal of Industrial and Production Engineering. 2022;39(5):399–413.

17. Dexter F, Ledolter J. Bayesian prediction bounds and comparisons of operating room times even for procedures with few or no historic data. The Journal of the American Society of Anesthesiologists. 2005;103(6):1259–167.

18. Eijkemans MJ, Van Houdenhoven M, Nguyen T, Boersma E, Steyerberg EW, Kazemier G. Predicting the unpredictable: a new prediction model for operating room times using individual characteristics and the surgeon’s estimate. The Journal of the American Society of Anesthesiologists. 2010;112(1):41–9.

19. Yuniartha DR, Masruroh NA, Herliansyah MK. An evaluation of a simple model for predicting surgery duration using a set of surgical procedure parameters. Informatics in Medicine Unlocked. 2021;25:100633. Available from: https://www.sciencedirect.com/science/article/pii/S2352914821001234.

20. Wang J, Cabrera J, Tsui KL, Guo H, Bakker M, Kostis JB. Clinical and Nonclinical Effects on Operative Duration: Evidence from a Database on Thoracic Surgery. J Healthc Eng. 2020 Feb;2020:3582796.

21. Abbas A, Mosseri J, Lex JR, Toor J, Ravi B, Khalil E, et al. Machine Learning using Preoperative Patient Factors Can Predict Duration of Surgery and Length of Stay for Total Knee Arthroplasty. International Journal of Medical Informatics. 2021 12;158:104670.

22. Marcon E, Kharraja S, Simonnet G. The operating theatre planning by the follow-up of the risk of no realization. International Journal of Production Economics. 2003 07;85:83–90.

23. Hans E, Wullink G, van Houdenhoven M, Kazemier G. Robust surgery loading. European Journal of Operational Research. 2008;185(3):1038–50. Available from: https://www.sciencedirect.com/science/article/pii/S0377221706005844.

24. Marques I, Captivo M, Pato M. An integer programming approach to elective surgery scheduling. OR Spectrum. 2012 04;34.

25. Shehadeh KS, Padman R. A distributionally robust optimization approach for stochastic elective surgery scheduling with limited intensive care unit capacity. European Journal of Operational Research. 2021;290(3):901–13. Available from: https://www.sciencedirect.com/science/article/pii/S0377221720307724.

26. American College of Surgeons. National Surgical Quality Improvement Program; 2021.

27. Liaw R, Liang E, Nishihara R, Moritz P, Gonzalez J, Stoica I. Tune: A Research Platform for Distributed Model Selection and Training. In: arXiv preprint; 2018.

28. Akiba T, Sano S, Yanase T, Ohta T, Koyama M. Optuna: A next-generation hyperparameter optimization framework. In: Proceedings of the 25th ACM SIGKDD international conference on knowledge discovery & data mining; 2019.

29. Ponce M, van Zon R, Northrup S, Gruner D, Chen J, Ertinaz F. Deploying a Top-100 Supercomputer for Large Parallel Workloads: The Niagara Supercomputer. In: Proceedings of the Practice and Experience in Advanced Research Computing on Rise of the Machines (Learning). New York, NY, USA: Association for Computing Machinery; 2019.

30. Rasmussen RV, Trick MA. The timetable constrained distance minimization problem. Annals of Operations Research. 2008 Jul;171(1):45. Available from: 10.1007/s10479-008-0384-4.

31. Laskin DM, Abubaker AO, Strauss RA. Accuracy of predicting the duration of a surgical operation. J Oral Maxillofac Surg. 2013 Feb;71(2):446–7.

32. Pandit JJ, Tavare A. Using mean duration and variation of procedure times to plan a list of surgical operations to fit into the scheduled list time. Eur J Anaesthesiol. 2011 Jul;28(7):493–501.

33. Pandit JJ. Rational planning of operating lists: a prospective comparison of ‘booking to the mean’ vs. ‘probabilistic case scheduling’ in urology. Anaesthesia. 2019 Dec;75(5):642–7.

34. Wang J, Cabrera J, Tsui KL, Guo H, Bakker M, Kostis JB. Predicting Surgery Duration from a New Perspective: Evaluation from a Database on Thoracic Surgery. arXiv; 2017. Available from: https://arxiv.org/abs/1712.07809.

35. Lai J, Huang CC, Liu SC, Huang JY, Cho DY, Yu J. Improving and Interpreting Surgical Case Duration Prediction with Machine Learning Methodology. medRxiv. 2020. Available from: https://www.medrxiv.org/content/early/2020/12/08/2020.06.10.20127910.

36. Rothstein DH, Raval MV. Operating room efficiency. Semin Pediatr Surg. 2018 Feb;27(2):79–85.

37. Hassanzadeh H, Boyle J, Khanna S, Biki B, Syed F. Daily surgery caseload prediction: towards improving operating theatre efficiency. BMC Medical Informatics and Decision Making. 2022 Jun;22(1):151. Available from: 10.1186/s12911-022-01893-8.

38. Strömblad CT, Baxter-King RG, Meisami A, Yee SJ, Levine MR, Ostrovsky A, et al. Effect of a Predictive Model on Planned Surgical Duration Accuracy, Patient Wait Time, and Use of Presurgical Resources: A Randomized Clinical Trial. JAMA Surgery. 2021 04;156(4):315–21. Available from: 10.1001/jamasurg.2020.6361.

39. Elmachtoub AN, Grigas P. Smart “Predict, then Optimize”. arXiv; 2017. Available from: https://arxiv.org/abs/1710.08005.

