## Supplementary Table 1 for "Machine Learning to Predict-Then-Optimize Elective Orthopaedic Surgery Scheduling Improves Operating Room Utilization"

**Supplementary Table 1.** Comparing two-stage, mean and hindsight duration of surgery using the Any schedule optimization formulation.

| Schedule Parameters | Mean Overtime, min/week (SD) | P-value | Mean Underutilization, min/week (SD) | P-value | Mean Cases/week (SD) | P-value |
|---|---|---|---|---|---|---|
| **Granularity = 10 min. Waitlist = 250** | | | | | | |
| Two-Stage | 1335.7 (322.1) | Reference | 669.1 (189.1) | Reference | 125.0 (0.2) | Reference |
| Mean | 1706.5 (372.1) | <0.001 | 655.7 (177.7) | 0.948 | 125.0 (0) | 0.083 |
| Hindsight | 0 (0) | <0.001 | 0 (0) | <0.001 | 117.2 (2.8) | <0.001 |
| **Granularity = 10 min. Waitlist = 500** | | | | | | |
| Two-Stage | 1410.4 (331.2) | Reference | 694.4 (157.0) | Reference | 125.0 (0.2) | Reference |
| Mean | 1728.1 (354.8) | <0.001 | 644.0 (178.4) | 0.011 | 125.0 (0) | 0.025 |
| Hindsight | 0.0 (0.0) | <0.001 | 0.0 (0.0) | <0.001 | 121.0 (4.5) | <0.001 |
| **Granularity = 10 min. Waitlist = 1000** | | | | | | |
| Two-Stage | 1368.8 (310.0) | Reference | 689.6 (183.7) | Reference | 126.9 (0.3) | Reference |
| Mean | 1804.0 (366.8) | <0.001 | 698.5 (180.9) | 0.745 | 126.9 (0.4) | 0.564 |
| Hindsight | 0.0 (0.0) | <0.001 | 0.8 (7.8) | <0.001 | 126.4 (5.8) | 0.764 |
| **Granularity = 10 min. Waitlist = 1500** | | | | | | |
| Two-Stage | 1267.9 (321.9) | Reference | 657.7 (165.2) | Reference | 126.8 (0.4) | Reference |
| Mean | 1452.4 (378.0) | <0.001 | 883.2 (287.2) | <0.001 | 123.0 (3.3) | <0.001 |
| Hindsight | 0.0 (0.0) | <0.001 | 0.0 (0.0) | <0.001 | 130.6 (4.1) | <0.001 |

| | | | | | | |
|---|---|---|---|---|---|---|
| **Granularity = 15 min. Waitlist = 250** | | | | | | |
| Two-Stage | 1416.2 (357.7) | Reference | 702.3 (185.5) | Reference | 123.4 (1.8) | Reference |
| Mean | 2001.3 (366.2) | <0.001 | 622.9 (176.1) | 0.007 | 125.0 (0.0) | <0.001 |
| Hindsight | 0.0 (0.0) | <0.001 | 0.0 (0.0) | <0.001 | 117.3 (2.7) | <0.001 |
| **Granularity = 15 min. Waitlist = 500** | | | | | | |
| Two-Stage | 1450.8 (331.3) | Reference | 674.4 (167.6) | Reference | 124.8 (0.4) | Reference |
| Mean | 2048.7 (364.7) | <0.001 | 661.6 (171.9) | 0.304 | 125.0 (0) | <0.001 |
| Hindsight | 0.0 (0.0) | <0.001 | 0.0 (0.0) | <0.001 | 121.0 (2.7) | <0.001 |
| **Granularity = 15 min. Waitlist = 1000** | | | | | | |
| Two-Stage | 1434.7 (324.7) | Reference | 669.7.5 (160.8) | Reference | 124.6 (0.7) | Reference |
| Mean | 2077.2 (396.5) | <0.001 | 663.6 (198.8) | 0.843 | 125.0 (0) | <0.001 |
| Hindsight | 0.0 (0.0) | <0.001 | 0.0 (0.0) | <0.001 | 122.6 (3.3) | <0.001 |
| **Granularity = 15 min. Waitlist = 1500** | | | | | | |
| Two-Stage | 1369.9 (356.1) | Reference | 641.3 (153.5) | Reference | 123.8 (1.2) | Reference |
| Mean | 2008.6 (370.0) | <0.001 | 599.4 (162.5) | 0.028 | 125.0 (0) | <0.001 |
| Hindsight | 0.0 (0.0) | <0.001 | 0.0 (0.0) | <0.001 | 125.2 (5.6) | 0.006 |
