## Supplementary Table 2 for "Machine Learning to Predict-Then-Optimize Elective Orthopaedic Surgery Scheduling Improves Operating Room Utilization"

**Supplementary Table 2.** Comparing two-stage, mean and hindsight duration of surgery using the Split schedule optimization formulation.

| Schedule Parameters | Mean Overtime, min/week (SD) | P-value | Mean Underutilization, min/week (SD) | P-value | Mean Cases/week (SD) | P-value |
|---|---|---|---|---|---|---|
| **Granularity = 10 min. Waitlist = 250** | | | | | | |
| Two-Stage | 1003.8 (238.1) | Reference | 285.2 (117.4) | Reference | 124.9 (0.2) | Reference |
| Mean | 1356.0 (277.4) | <0.001 | 194.9 (94.1) | <0.001 | 125.0 (0.0) | 0.046 |
| Hindsight | 0.0 (0.0) | <0.001 | 0.0 (0.0) | <0.001 | 116.1 (3.0) | <0.001 |
| **Granularity = 10 min. Waitlist = 500** | | | | | | |
| Two-Stage | 966.7 (232.9) | Reference | 300.8 (113.9) | Reference | 125.0 (0.1) | Reference |
| Mean | 1253.3 (277.9) | <0.001 | 212.5 (92.0) | <0.001 | 125.0 (0) | 0.317 |
| Hindsight | 0.0 (0.0) | <0.001 | 0.0 (0.0) | <0.001 | 120.5 (3.3) | <0.001 |
| **Granularity = 10 min. Waitlist = 1000** | | | | | | |
| Two-Stage | 964.0 (243.9) | Reference | 301.6 (119.9) | Reference | 125.0 (0.2) | Reference |
| Mean | 1319.7 (289.7) | <0.001 | 216.9 (106.8) | <0.001 | 125.0 (0) | 0.025 |
| Hindsight | 0.9 (8.8) | <0.001 | 5.4 (55.6) | <0.001 | 119.9 (3.9) | <0.001 |
| **Granularity = 10 min. Waitlist = 1500** | | | | | | |
| Two-Stage | 972.2 (225.2) | Reference | 325.9 (114.8) | Reference | 124.9 (0.3) | Reference |
| Mean | 1258.9 (265.6) | <0.001 | 335.1 (241.6) | 0.497 | 123.8 (2.4) | <0.001 |
| Hindsight | 12.8 (20.7) | <0.001 | 176.6 (248.5) | <0.001 | 116.3 (5.3) | <0.001 |

| | | | | | | |
|---|---|---|---|---|---|---|
| **Granularity = 15 min. Waitlist = 250** | | | | | | |
| Two-Stage | 985.1 (235.6) | Reference | 307.6 (119.9) | Reference | 123.1 (1.9) | Reference |
| Mean | 1497.3 (308.5) | <0.001 | 156.3 (78.7) | <0.001 | 125.0 (0.0) | <0.001 |
| Hindsight | 0.0 (0.0) | <0.001 | 0.0 (0.0) | <0.001 | 114.5 (2.9) | <0.001 |
| **Granularity = 15 min. Waitlist = 500** | | | | | | |
| Two-Stage | 1016.3 (240.1) | Reference | 273.9 (104.0) | Reference | 124.9 (0.3) | Reference |
| Mean | 1516.7 (228.9) | <0.001 | 153.9 (84.1) | <0.001 | 125.0 (0) | <0.001 |
| Hindsight | 0.0 (0.0) | <0.001 | 0.0 (0.0) | <0.001 | 116.6 (5.4) | <0.001 |
| **Granularity = 15 min. Waitlist = 1000** | | | | | | |
| Two-Stage | 1039.3 (229.5) | Reference | 242.0 (99.3) | Reference | 124.9 (0.4) | Reference |
| Mean | 1538.8 (272.9) | <0.001 | 139.0 (71.2) | <0.001 | 125.0 (0) | <0.001 |
| Hindsight | 0.0 (0.0) | <0.001 | 0.0 (0.0) | <0.001 | 117.8 (5.4) | <0.001 |
| **Granularity = 15 min. Waitlist = 1500** | | | | | | |
| Two-Stage | 996.2 (207.1) | Reference | 294.1 (108.3) | Reference | 124.5 (0.7) | Reference |
| Mean | 1531.9 (245.3) | <0.001 | 149.9 (71.9) | <0.001 | 125.0 (0) | <0.001 |
| Hindsight | 3.6 (36.6) | <0.001 | 0.0 (0.0) | <0.001 | 120.5 (4.3) | <0.001 |
